# Teleconferencing as an alternative to written Advice and Guidance referrals at the primary-secondary care interface: a qualitative case study

**DOI:** 10.64898/2026.02.12.26343579

**Authors:** Shan He, Juliet A Usher-Smith, Graham Martin

## Abstract

**Background:** Communication issues across the primary-secondary care interface are considered one of the most important challenges in improving patient safety in primary care in the UK. Teleconferencing offers a potential means of improving communication during referrals but is largely unevaluated.

**Aim:** To explore teleconferencing as an alternative to written Advice and Guidance (A&G) referrals for neurology cases, by assessing its impact on GP-specialist communication and relationships, and exploring implications for patient care.

**Design and Setting:** A qualitative case study of a primary care network (PCN) and a secondary care centre in East Anglia.

**Methods:** 18 clinicians and 10 other stakeholders were interviewed. Observations of teleconferences and a focus group with five PCN staff provided additional data. Data collection and analysis were guided by the Consolidated Framework for Implementation Research and Reflexive Thematic Analysis.

**Results:** Advantages of teleconferencing identified by participants included greater clinician satisfaction, mutual educational value, streamlined patient journeys and continuity of care. Teleconferences were also seen to build GP-specialist relationships and reduce unnecessary outpatient referrals. Perceived issues included time constraints, clinical governance and funding sustainability; teleconferences were not seen as appropriate for all referrals. Overall, participants welcomed the teleconference approach but stressed the need to robustly assess its cost-effectiveness and replicability in other settings.

**Conclusion:** Teleconferencing is a potentially promising alternative to written A&G referrals and was perceived by participants to help build GP-specialist relationships. However, further studies are needed to assess clinical effectiveness and costs, and to guide future development and implementation.

**How this fits in:** *What is known?:* Referral interventions involving direct GP-specialist dialogue can enhance referral quality, reduce outpatient referrals and improve GP-specialist relationships, with some demonstrating improved clinical outcomes. However, they often face sustainability challenges, and their cost-effectiveness and mechanisms of impact require further assessment.

*What does this study add?:* This qualitative study identifies key mechanisms through which virtual GP-specialist dialogue may lead to downstream benefits: enabling shared decision-making and delivering consultant-level care closer to home; empowering GPs to manage complex cases; and reducing overall workload across primary and secondary care systems. The programme theory developed can be used to guide future intervention design, implementation and evaluation.

## Introduction

The UK’s James Lind Alliance Priority-Setting Partnership identified communication issues across the primary-secondary care interface as a key research priority for patient safety in primary care.^1^ Problems with the interface consume around 15 million general practitioner (GP) appointments annually, adversely affecting patient experience, safety and workload.^2^

With escalating outpatient referrals and long waiting times, expansion of Advice & Guidance (A&G) in England has helped slow GP referral rates to consultants.^3^ A&G allows GPs to access specialist advice before or instead of formal referrals, through synchronous (e.g., telephone) or asynchronous (e.g., written e-referral) methods.^4–6^ While intended to promote shared decision-making and avoid unnecessary outpatient activities,^5^ GPs have raised concerns that written A&G shifts workload to primary care and risks becoming a mandatory gateway, blocking direct outpatient referrals.^7^ This shift was recognised in the 2025/2026 GP contract, which allocated £80m to support primary care engagement with A&G.^8,9^

Referral interventions involving direct GP-specialist dialogue can enhance referral quality, reduce outpatient referrals and improve GP-specialist relationships, with some demonstrating improved clinical outcomes. However, they often face sustainability challenges, and their mechanisms of impact remain poorly understood.^10–14^ Teleconferences between GPs and specialists have shown benefits in chronic disease management, reducing referrals and in the case of diabetes management, improving clinical outcomes.^10,15^ However, evidence of their use for neurology referrals is limited.

This study examines a neurology teleconferencing initiative in the East of England, exploring whether synchronous dialogue can overcome challenges associated with written A&G. It formatively evaluates its impact on GP-specialist relationships, clinician satisfaction, workload, and implications for patient care, while developing a programme theory of how teleconferencing exerts its impact.

## Methods

This qualitative case study was conducted in an acute trust and a Primary Care Network (PCN) comprising six practices in East Anglia. Interview questions were shaped by the Consolidated Framework for Implementation Research^16^ and refined by pilot interviews with the lead neurologist and the lead GP, conducted at the pre-funding stage. Research Ethics Committee and Health Research Authority (HRA) approvals were obtained (IRAS: 325193, REC ref: 23/NE/0111).

### Theoretical approach

Following MRC guidance,^17–19^ the study aimed to understand how teleconferencing works by developing a preliminary programme theory, identifying its “active ingredients” and the mechanisms of influence on outcomes and experiences of service users, clinicians and the healthcare system. This approach captured emerging changes in implementation, experiences of the intervention and unanticipated pathways, to inform future feasibility and effectiveness evaluations.^17^

### The intervention

Initiated in early 2020 by clinicians, the teleconferencing intervention aims to improve the quality of communication relating to neurology referrals. It involves fortnightly one-hour teleconferences connecting PCN GPs with a consultant neurologist via Microsoft Teams, typically covering 10-15 patients. Patients are referred by individual GPs via electronic messaging within SystmOne.

Teleconferences are led by a GP and neurologist and designed to be informal and flexible, with referring GPs encouraged to “present and leave” to facilitate participation. Most cases require advice, reassurance, direct investigations, or redirection to more appropriate specialties. When formal referrals are necessary, referring GPs write brief referral letters and the neurologist assigns triage codes to bypass standard triage.

Outside teleconferences, GPs can contact the neurologist via Teams chat and arrange ad-hoc calls. Educational materials, e.g. headache guidelines, are made available to the participating PCN as part of the intervention.

### Data collection

Data were collected from September 2023 to May 2024 through semi-structured interviews, a focus group discussion and observations of five teleconferences (six hours). Participants were recruited purposively using snowball sampling. Data analysis occurred alongside data collection. Recruitment ceased when theoretical saturation was reached, i.e., no new themes or insights emerged from additional interviews.

### Data management and analysis

The study drew on reflexive thematic analysis, acknowledging the lead researcher’s GP status and how that might influence data collection and interpretation.^20^ Two researchers independently coded the first four interviews to establish consensus on the initial codes and themes. The lead researcher then coded the remaining data in NVivo 14. Coding was largely inductive, with CFIR-based deductive codes added later to deepen the analysis.^16^ We compared themes across interviews, focus group data and observations, examining where accounts converged or diverged across stakeholder groups.

Emerging themes were discussed regularly with the research team to refine codes and themes,^21^ and with the PCN Patient Participation Group (PPG) to ensure that analysis was informed by patient experiences.^22^

## Results

### Study participants

Twenty-eight individuals participated, including 18 clinicians and administrative staff across primary and secondary care and 10 other stakeholders from the integrated care system (ICS), Local Medical Committee (LMC) and hospital (see Table 1).

**Table 1:** Characteristics of study participants (Note: one participant contributed to both the interview and the focus group discussion. Some participants had dual roles. The primary role for which they were recruited is listed.)

|  | Participants | Number |
| --- | --- | --- |
| Interviews – clinicians | GPs | 6 (4 Partner GPs and 2 Salaried GPs) |
|  | Neurologists | 5 |
|  | Psychiatrists | 2 |
|  | Advanced Nurse Practitioners | 2 |
| Interviews – other stakeholders | ICS leaders (GPs) | 3 |
|  | Hospital managers | 3 |
|  | LMC leader (GP) | 1 |
|  | Hospital/PCN secretaries | 2 |
| Focus group discussion | GPs | 4 (All Partner GPs) |
|  | PCN secretary | 1 |

### Overview of results

The dataset provided insights into four overarching themes: the context for the primary-secondary care referral interface; clinicians and wider stakeholders’ attitudes towards teleconferencing; GP-specialist relationship-building and its system-level implications; and issues with teleconferences. A preliminary logic model (Figure 1) was developed iteratively through reflexive thematic analysis, prior literature review and stakeholder validation. ^23^

**Figure 1.**
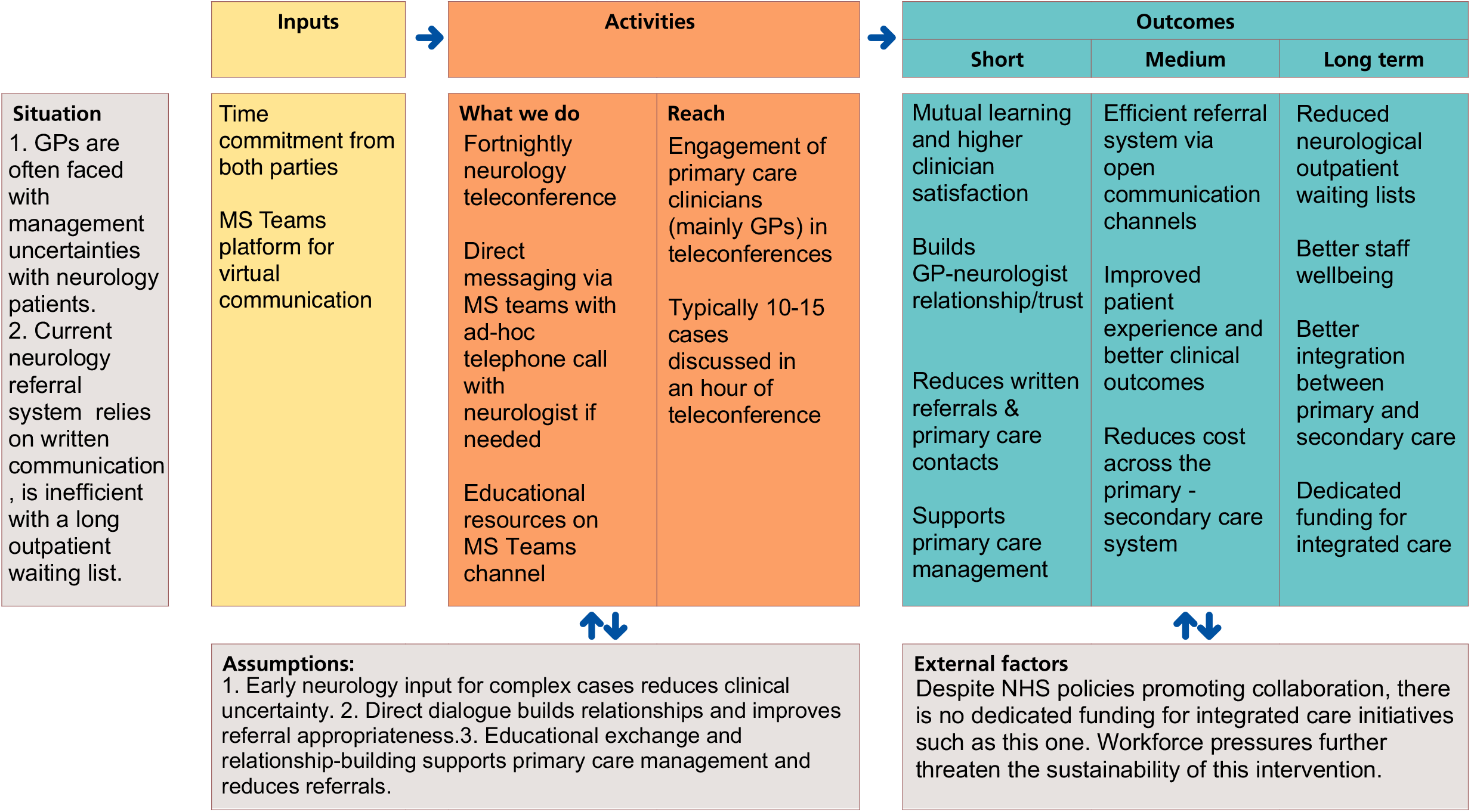
**Logic model** underlying neurology teleconferencing referral intervention. A diagrammatic roadmap illustrating the pathway from direct dialogue to system-level impacts across the primary-secondary care interface.

#### 1. The context for referrals across the primary-secondary care interface

##### 1.1 Faceless interface and increasing tension

Since routine referrals occurred mostly via letters (written A&G or outpatient referrals), participants noted GPs and specialists rarely spoke except for urgent referrals. This “faceless” interface, as multiple participants put it, created tensions, exacerbated by strained capacity in both services:

> *“Now, when everyone is particularly stressed and frustrated, I think the tone of the letters is getting more terse…people are a lot more unpleasant to each other when they don’t have a face to go with it.” [Neurologist-P7]*

This lack of direct interaction resulted in patients “bouncing” between systems with no evident coordination.

> *“The relationship probably broke down quite a lot [during the Covid-19 pandemic]…there’s a lot of pushing backwards and forwards…they are annoyed with each other quite clearly…sometimes the patient gets lost in that.” [Hospital manager-P13]*

##### 1.2 Perceived workload transfer from secondary to primary care

GPs and other stakeholders perceived inappropriate workload transfer from secondary to primary care due to long outpatient waiting lists and tightened referral thresholds, with increasing numbers of referrals rejected or re-directed:

> *“If you’re waiting a year to see a specialist, you don’t just sit at home waiting, you will call your GP lots of times in the meantime, because your problem is probably getting worse. If your specialist then offers you a phone appointment, and you feel that that hasn’t answered all your questions, you’ll go back to your GP.” [LMC/GP-P20]*

Moreover, primary care clinicians voiced concerns about the to-do lists generated by written A&G responses:

> *“[Written A&G] is particularly like that…And they’ll often just ping it back, and they’re quite patronising…they come back with almost making you do the work.” [ANP-P3]*

GPs responded to workload shifting by, for example, ‘working to rule’ by using the British Medical Association’s template letters:^24^

*“We’re under so much pressure that we’re putting our foot down…everyone’s gone back to the contract…arguably from a patient perspective it’s better we do it. But when we’re not paid or not contracted, we’re going to push back, we haven’t got the workforce.” [ICS/GP-P21]*

In an increasingly protocol-driven referral system, clinicians on both sides perceived a degree of ‘gaming’ of referrals to get them accepted (e.g., by ticking a box):

> *“GPs are very clever and they’ve adapted the way that they refer…So now what we’re getting is a lot of very limited, ‘I’m concerned about this patient, she has refractory headaches’.” [Neurologist-P7]*

Interestingly, views on referral proformas diverged: ICS leaders valued them for clinical governance, whereas GPs felt rigid proformas could not accommodate complexity and might encourage ‘gaming’ of referrals. One ICS leader suggested conversations alongside written referrals to mitigate these risks.

Tension across the interface was worsened by unclear responsibilities, e.g. for incidental findings. Despite attempts by the ICS, LMC and hospital managers to develop interface principles, resolution typically required case-by-case discussions. Many participants suggested that direct clinician-to-clinician conversations could help build relationships across the primary-secondary care interface and improve patient care.

#### 2. Clinicians and wider stakeholders’ attitudes towards teleconferencing

##### 2.1 Enhanced communication

Clinicians on both sides of the interface welcomed A&G via teleconferencing for the way, in contrast to written A&G, it enabled nuanced, two-way communication, allowing the expression of patients’ and GPs’ ideas, concerns and expectations (ICE).^25^ This was particularly valuable for ‘grey’ or ‘middle-ground’ queries, where management options were not clear-cut.

> *“The ones in the middle, where it is less certain, I think that’s where the dialogue is particularly useful…Which option one chooses is largely down to…understanding more about the patient and understanding more about the clinician, the GP.” [GP-P4]*

Both GPs and neurologists contrasted the quality of communication they experienced during teleconferences with that of written A&G. The lead neurologist explained how he was better able to address GPs’ ICE, facilitated by relationship-building:

> *“If [GP] calls me, I can tell from how he asks the question what he wants me to do. Whereas that’s not there in advice and guidance, right*… *In the same way that the way a GP’s talking to me, I can tell what the problem is and what they need and what they want, right…and just by knowing people as well.” [Neurologist-P6]*

With written A&G, conversely, neurologists sometimes did not understand questions asked in letters; GPs reported that questions remained unanswered following outpatient appointments. Resolving communication issues could require multiple letter exchanges.

> *“[With written A&G,] you send a single message, and you get a message back, and it might not be that informative and actually it might be very contrary to what you thought they were going to say, and it’s not always helpful. And then you have to send another A&G and, you know, three, four, five weeks pass.” [FG-P4]*

A&G letters were considered ‘transactional’ and suitable only for straightforward scenarios. In contrast, teleconferences were considered useful for grey areas and complex case management, e.g., patients with functional neurological disorders.

##### 2.2 Mutual educational benefits

Clinicians reported that teleconferences facilitated mutual clinical and organisational learning. GPs gained confidence in managing neurological conditions:

> *“A lot of us are much more confident now, if something’s been discussed in the meeting several times…you might try some treatments or you know what the answer is.” [GP-P1]*

Neurologists gained insight into the challenges of working in primary care, such as undifferentiated initial patient presentations and limited resources, prompting them to consider the impact of their advice on primary care workload. This mutual understanding enabled negotiation of appropriate thresholds for referrals.

> *“So we get to know what’s turning up at primary care. But also…maybe appreciating where the level of risk or management is and where to set that.” [Neurologist-P15]*

Participants also saw teleconferencing as a way of facilitating broader learning by identifying and addressing underlying pathway or process issues:

> *“What we also encourage from this is that wider learning…if the problem identifies something in the pathway or the process or something… that we can actually resolve the underlying problem.” [ICS/GP-P14]*

##### 2.3 Higher clinician satisfaction

Clinicians on both sides expressed satisfaction with neurology teleconferences:

> *“I really, really like it and actually I think it just reminds you that everyone is trying to do their best for their patients.” [Neurologist-P7]*
>
> *“In a time when professional burnout and morale is at an all-time peak, relationship-based care with consultant colleagues is also a powerful antidote.” [GP-P4]*

Their positive views stemmed from perceived benefits for patients, clinicians and the system.

> *“If actually the integration was based upon the mutual agreement that we want the patient journey to be the least convoluted, we want the best outcome for the patient, then working together should be seen as a massive positive. Because every time that specialist works with us we learn…And the more we interact with specialists the more they should be able to see the benefit.” [FG-P3]*

#### 3. Relationship-building through teleconferences

Central to the value GPs and specialists identified in teleconferencing was that it fostered goodwill and trusting relationships, with potential benefits for patient care.

> *“The consultant trusts us, we trust him. When you pick up the phone and say, ‘I’m really worried’, he’s built a relationship and understanding of you and goes, ‘If you’re worried, I believe you, I’m going to do this’. That patient care is priceless.” [FG-P3]*

In contrast, conventional written referrals offered little scope for relationship-building.

> *“In general, with the neurologists, I have no real knowledge about them as individuals or their styles or their approach, so it feels distant, remote, impersonal and frequently transactional.” [GP-P4]*

Both parties preferred ‘old’ ways of making referrals, predating written A&G, where telephone contact was more common. They viewed teleconferencing as a way of rebuilding primary-secondary care relationships.

> *“We’re so stuck in bureaucracy and processes…dare I say it, for medicolegal reasons really, that we’re less able to be informal and do things which then enhance the relationship.” [ICS-P21]*

Participants argued that these improved relationships could strengthen primary care, reduce workload across primary-secondary care systems, and improve patient care.

##### 3.1 Strengthening primary care

Participants suggested that stronger relationships with consultants could empower GPs to hold more clinical risk and manage more complex patient cases, thereby strengthening primary care’s role in the broader healthcare system.^26^ However, they acknowledged that not all GP practices would be comfortable with this role without additional funding.

> *“We’re taking more off the consultants. So, it’s vitally important that that is understood because we are containing things within our own basket of many things to carry, with a nice relationship with the consultant that we trust, who we know is going to support and help us.” [FG-P3]*

This enhanced gatekeeping was facilitated by quick access to specialist advice, both during teleconferences and via Teams chat, avoiding unnecessary outpatient referrals.

> *“Having access to advice around the time of a referral is always useful… It’s very hard to get advice and sometimes we just have to refer because…we can’t get an answer*.” (LMC/GP-P20)

In one example, access to the neurologist helped avoid an ED attendance for a complex patient with known diagnosis of myasthenia gravis by spotting early signs of deterioration and admitting him directly under neurology. This early treatment allowed a shorter hospital stay. Following discharge, close liaison between the two parties supported medication titration and clinical monitoring in the community.

##### 3.2 Reducing workload across primary-secondary care systems

Both GPs and neurologists agreed that, compared with written A&G, teleconferencing was workload neutral at an individual level but reduced overall workload across the primary-secondary care system. Relationship-building between GPs and specialists facilitated open dialogue and appeared central to these system-level benefits.

For primary care, workload reductions resulted from fewer written referrals, significantly reduced administrative burden, and fewer follow-up appointments:

> *“You’re getting information from the specialist quickly, which stops the patients bouncing around and coming back, ‘I’ve still got this, you haven’t fixed this, I need more pain relief’.” [GP-P5]*

For secondary care, reduced workload mainly arose from better triage and reduced referrals.

> *“The advantages are that it reintroduces that personal one-to-one relationship with consultant colleagues. It allows for a free and open expression of doubt and uncertainty as well as conveying certainties, and it can be useful in avoiding referrals when the primary purpose of the debate is to provide consultant-level reassurance to the GP.” [GP-P4]*

Written A&G, in contrast, was less effective as a triage tool (e.g., sicker patients sometimes being referred on a slower pathway). Rejecting referrals was time-consuming and created job dissatisfaction amongst neurologists. Additionally, many rejected referrals were later re-referred, creating additional inefficiencies. One neurologist voiced his frustration with the workload generated by written A&G:

> *“Advice and Guidance, I can see why it’s being pushed, but I think there’s a limit and the workload now for Advice and Guidance is huge…And I can’t help feeling it’s a way of covering people’s backs to a certain extent.” [Neurologist-P15]*

##### 3.3 Improvements to patient care

The continuity of advice from the same neurologist to GPs brought about by teleconferencing was widely perceived to bring patient benefits including effective reassurance, streamlined patient journeys, and shared decision-making, with PPG feedback supporting these views.

Teleconferences allowed GPs to consult a neurologist about patients’ concerns, providing more effective reassurance:

> *“I can turn around and say, ‘I can talk to a consultant neurologist about this and then I’ll be back in touch with you’. And that in itself carries power, especially when you’ve got a frequent attender who’s anxious.” [FG-P4]*

Teleconferencing was also seen to streamline patient journeys from initial presentation to appropriate diagnosis and treatment, reducing unnecessary steps in the care pathway. Patients appeared to move faster through the referral process thanks to rapid neurological opinion, direct access to investigations, and appropriate triaging to sub-specialties:

> *“[It’s] the perfect scenario, I’ve got very rapidly the opinion I wanted, I’ve got the investigation I’m not allowed to sort, my patient is going to get sorted a lot quicker, they don’t need to see the neurologist, I’ve seen and examined them. Now if their MRI is normal, they will get a nice letter from them and if their MRI is abnormal, they will get pulled into the neurology clinic.” [GP-P2]*

Similarly, GPs and neurologists reported that relationship-building enabled shared decision-making, delivering consultant-level care closer to home, with potential benefits for patient safety and continuity.

> *“It’s continuity with specialty. And that builds relationships, and that changes culture. And changing culture saves money and time and improves the patient journey.” [FG-P3]*
>
> *“He’s co-owning the responsibility…I think by sharing it, as you would with any case in general practice, you would ask a colleague or a colleague would ask you, you’d share the decision-making. It does make things, I think, safer.” [ANP-P3]*

#### 4. Issues with teleconferencing

Though participants were positive about teleconferencing, they noted limitations and opportunities for improvement.

##### 4.1 Time constraints

The synchronous nature of teleconferences meant that GPs often struggled to attend meetings due to part-time working or competing demands. In these cases, the lead GP would present on their behalf, with follow-up from the neurologist to the referring GP.

> *“Sometimes it feels…a hassle to try and get to it because you’ve got lots of other things…and it feels a bit, ‘I’ve got to do this now’. But that’s more just pressure of being a GP.” [GP-P9]*

##### 4.2 IT and Information governance

One GP highlighted that neurologists had limited access to SystmOne, the primary care electronic patient record system, and so relied on GPs to present cases. Additionally, teleconference referrals did not free GPs from all administrative steps, with outpatient referral letters still required afterwards. It was suggested that a dedicated administrator could summarise discussions and send referrals directly to neurology.

Neurologists raised information governance concerns regarding data-hosting, and noted that verbal communication could be misinterpreted, though efforts were underway to address these issues. While “transactional”, written A&G did at least provide an audit trail and medico-legal protection for clinicians.

##### 4.3 Funding sustainability

Teleconferences were initially unfunded, relying on clinicians’ goodwill. The acute trust later granted one-year funding for teleconferences at the pilot PCN and a neighbouring PCN to assess cost-effectiveness, though long-term sustainability remained uncertain. One hospital manager highlighted misaligned financial incentives: the acute trust voluntarily funded an intervention that might reduce its income by reducing outpatient referrals, while benefits accrued across the wider system.

Despite the introduction of ICSs, there was no dedicated funding for joined-up initiatives like this. One ICS leader suggested that money could be shifted from secondary to primary care while acknowledging the challenges:

> *“Now the challenge we have is actually if the benefit is felt elsewhere in the system by another bit of the system, how do you get that money in? But that’s another game.” [ICS/GP - P14]*

##### 4.4 Use as a mandatory gateway for all referrals

Driven by funding constraints and the need to demonstrate cost savings, teleconferences became the mandated referral pathway for the two PCNs, replacing both written A&G and direct outpatient referrals. Observation revealed the impact: teleconferences increased from an average of 1h to 1.5h due to increased volume of cases.

This shift proved controversial. GPs, including ICS and LMC GP leaders, and neurologists cautioned against mandatory gateways, arguing clinicians should have direct access to outpatient referrals. They highlighted that teleconferences were not appropriate and lacked capacity for all referrals:

> *“The concern I have around those systems is if…everything has to go through that system. So there’ll be some patients…I just want to get a referral in, I know they need to be seen. You wouldn’t want to be an artificial block in that system.” [*LMC/GP-P20]

One salaried GP described needing to discuss a patient with suspected Parkinson’s disease via teleconferencing simply to “rubber stamp” the referral. In contrast, several partner GPs supported the mandatory gateway to secure investment and enable nuanced case discussions that might avoid outpatient referrals. Some suggested that teleconferences should be reserved for ‘grey’ cases, with straightforward cases following standard pathways.

##### 4.5 Questions around cost-effectiveness and replicability

Many participants felt teleconferences would be cost-effective “in the long run” by reducing referrals but emphasised the need for robust evaluation.

> *“I think it’s definitely worth our time in the long run…Even if it is for a closer working relationship, managing patient and GP satisfaction with the service, decreasing their frustration…managing patients’ expectations as well and getting them to be treated in a timely manner in the most appropriate places.” [Neurologist-P27]*

While participants recommended scaling up the teleconferencing intervention, they questioned how well it would work beyond the pilot PCN or beyond neurology. They noted the unique characteristics of the PCN, with fully merged management and IT systems, GP and neurologist champions, and the peculiarities of neurology as a speciality with many “grey areas” as key in determining the success of the intervention. They recommended an incremental implementation strategy tailored to local needs, resources and each specialty. Table 2 summarises the key differences between teleconferencing and written A&G referrals as perceived by participants.

**Table 2:**
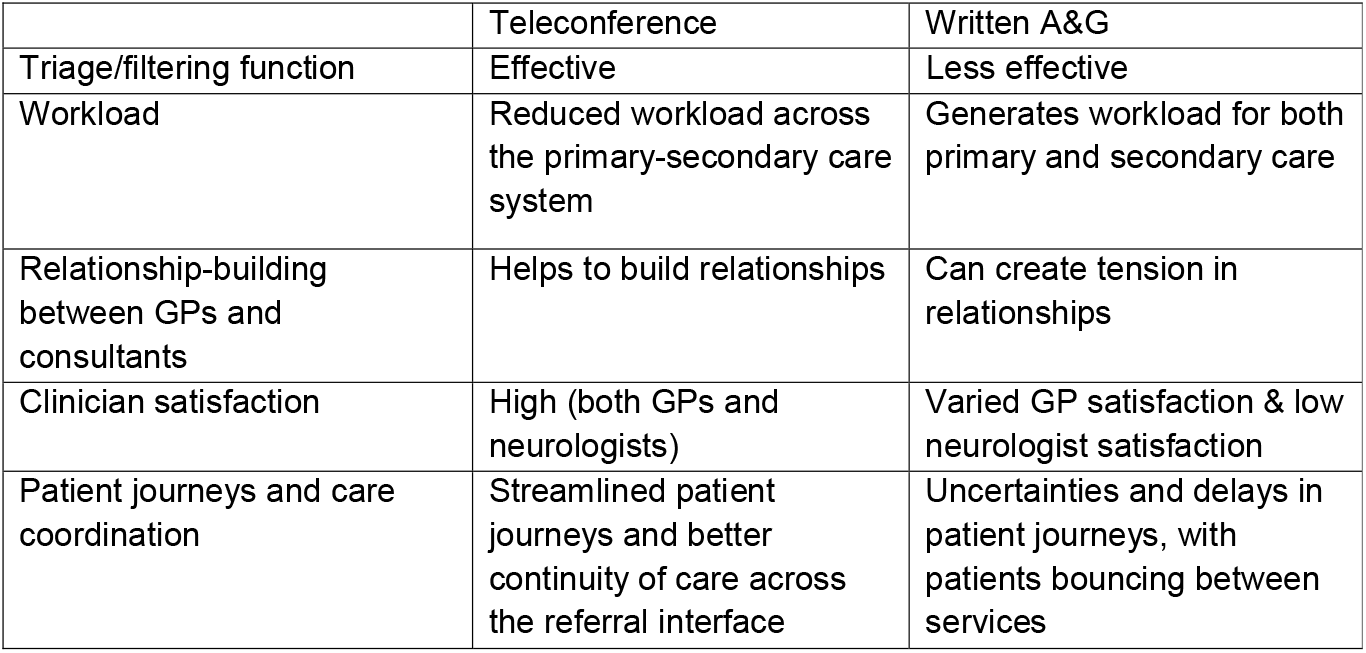
Comparison of teleconferencing with written A&G referrals across key dimensions.

## Discussion

### Summary

Participants reported that, compared with written A&G, teleconferences enhanced communication quality, provided mutual educational benefits, and improved clinicians’ satisfaction. Teleconferences helped build GP-neurologist relationships across an increasingly ‘transactional’ and strained interface, with potential benefits for patients, clinicians and the healthcare system. Challenges included time constraints, governance issues, funding sustainability, and concerns about teleconferences becoming mandatory gateways.

This analysis informed a mechanisms of impact map (Figure 2), which identifies building GP-specialist relationships and trust as the “active ingredient”. The logic model and mechanisms of impact map together illustrate how the intervention may strengthen primary care, reduce workload at system level, and foster better continuity across the interface.

**Figure 2.**
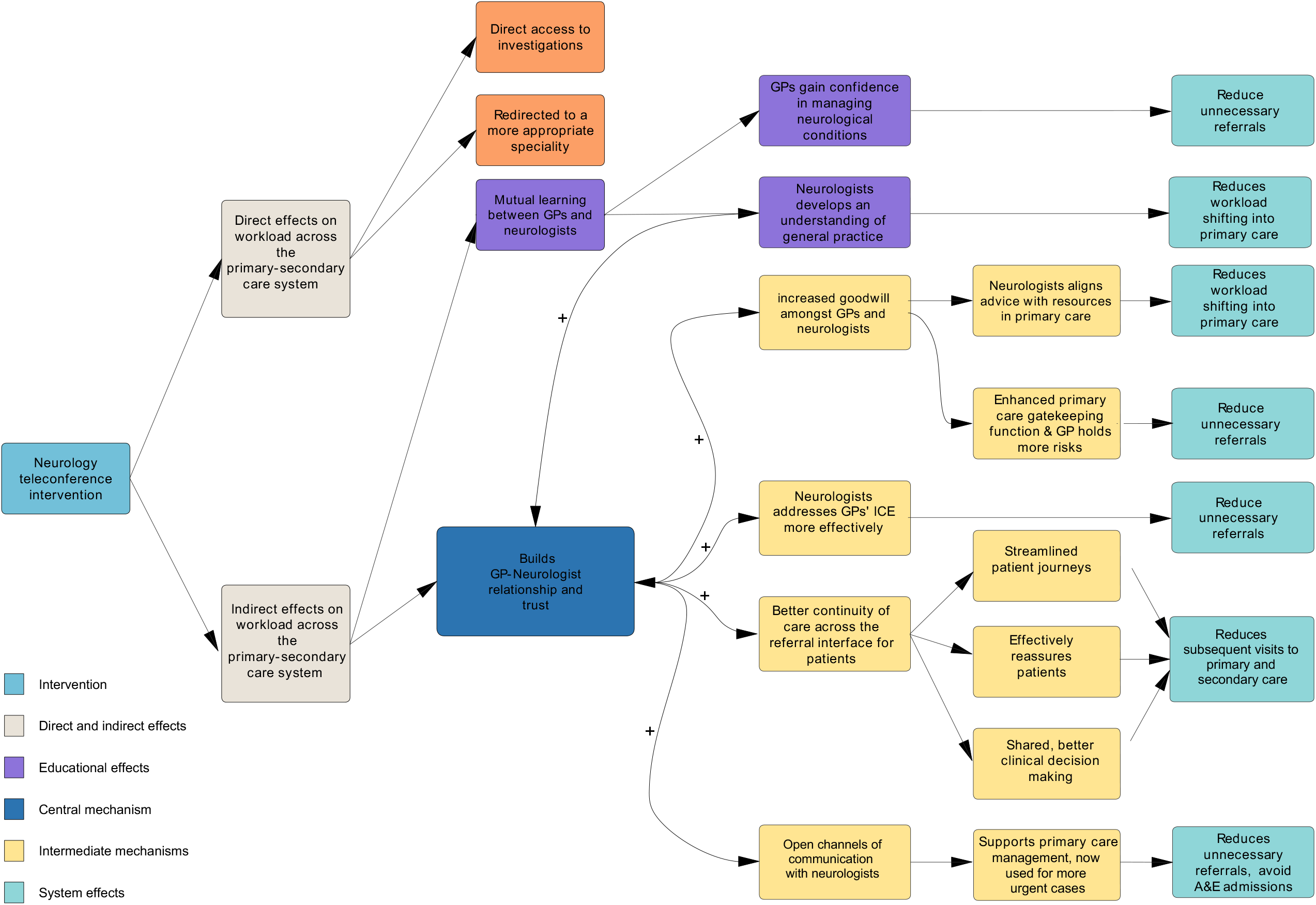
**Mechanisms of impact map** underlying neurology teleconferencing intervention. A diagrammatic representation showing how relationship-building between GPs and specialists acts as the active ingredient, generating cascading benefits at patient, clinician, organisational and system levels. The “+” signs indicate positive feedback loops that reinforce relationships and trust over time.

### Strengths and limitations

The qualitative case study approach enabled exploration of teleconferences in real-world settings and identified potential causal mechanisms through which teleconferences might achieve system-level impacts.^19,27^ Observation of teleconferences allowed further understanding of social and clinical context.

The inclusion of clinical and non-clinical participants from primary and secondary care ensured the study accounted for multiple perspectives. Data collection approximately three years post-implementation captured the evolving nature of the intervention, including its adaptation to external changes and influences.

The lead researcher’s status as a local GP facilitated recruitment and in-depth understanding of participants’ experiences but risked interpretive bias. This was mitigated through reflexive field notes and regular team discussions that challenged emerging interpretations. Moreover, Partner GPs were over-represented, whose views may differ from those of salaried GPs. While the PCN PPG contributed to interpretation of study findings, patients were not interviewed. This study does not assess safety, effectiveness or cost-effectiveness.

### Comparison with existing literature

Communication difficulties between primary and secondary care in the UK, including workload dumping and ‘operational failures’, are well documented and negatively affect GP-specialist relationships and patient care.^28–30^ Previous studies show that referral interventions that improve dialogue between GPs and specialists can enhance referral quality, reduce outpatient referrals and improve GP-specialist relationships, with some demonstrating improved clinical outcomes. However, they often face sustainability challenges and their cost-effectiveness requires further assessment.^10–14^ Our study adds to this evidence base by demonstrating mechanisms through which better GP-specialist relationships may benefit clinicians, patients and healthcare systems. In line with Nugus et al.’s study in emergency departments, which found referral interfaces to be porous and negotiable, our study suggests that navigating referral processes is about managing those boundaries through better communication and relationships, facilitating effective care coordination.^31^

Electronic means of referrals, specifically, have been explored in the UK.^32–35^ Although email communication allowed quick answers, clinicians in one study felt that it was not a firm basis for building relationships.^33^ Wallace et al.’s RCT of joint teleconsultations showed increased patient satisfaction but the costs of clinicians’ time were high, suggesting that certain specialities may be more suitable and that appropriate patient selection is crucial to cost-effectiveness.^34,35^ Our study offers some insight into the areas where interventions of this kind might plausibly improve quality while reducing costs, demonstrating how teleconferencing is particularly relevant to what participants called “grey areas”, common in disciplines like neurology. Other comparable interventions internationally have achieved impressive impacts. For example, heart failure virtual clinics in Ireland drastically reduced outpatient referrals (9% vs 93%) while benefiting underserved populations.^15^

Continuity of care, a key primary care quality domain,^36–39^ is associated with lower use of out-of-hours services, fewer acute hospital admissions, and lower mortality.^40^ It is defined by Haggerty et al. as how patients experience integration of services and coordination over time, comprising relational, informational, and management continuity.^41^ Teleconferencing appears to enhance informational continuity (efficient transfer of clinical and psychosocial information), and management continuity (responsive, shared management plans) across the referral interface. Additionally, teleconferencing improves GP-specialist relational continuity, aligning with Ladds and Greenhalgh’s concept of ‘continuity of distributed work’ between providers.^38^ The enhanced GP-specialist continuity may operate through similar mechanisms to GP-patient relational continuity: by enhancing open dialogue, efficient information transfer, greater GP responsibility, and more responsive specialist input, resulting in better clinician decision-making and care coordination.^41,42^ This relationship-based approach is particularly relevant as UK primary care becomes increasingly fragmented and transactional post-COVID-19.^43,44^

### Implications for policy and practice

Our findings suggest that teleconferencing is a promising intervention with potential benefits for both systems through improved coordination and workload reduction. Future investment in referral pathways between primary and secondary care should prioritise interventions that build relationships between services and support innovative, value-driven, preventative solutions.

Further evaluation might examine patients’ experiences, effectiveness and cost-effectiveness, identify which patient groups benefit most, and assess transferability to other specialties and regions. If scale-up proves feasible, such relationship-based referral models may strengthen primary care, support integrated care and the policy goal of shifting care from hospital to community^45^, with potential to ease pressure on outpatient services.

## Data Availability

All data produced in the present study are available upon reasonable request to the authors.

## Acknowledgements

We thank the Patient Participation Group for the case study Primary Care Network for their support and contribution in interpreting research findings. We thank the research participants who gave their time generously. We thank Dr Tim Wright and Dr Nushan Gunawardana for their input into the research and feedback on a draft of this article.

## Ethical approval

Ethical approval was obtained from North East - Tyne & Wear South Research Ethics Committee (June 2023, IRAS: 325193, REC ref: 23/NE/0111).

